# Biallelic *PAX7* variants cause a novel Satellite Cell-opathy with progressive muscle involvement resembling facioscapulohumeral muscular dystrophy

**DOI:** 10.1101/2025.03.03.25322917

**Authors:** Massimo Ganassi, Claudia Strafella, Marco Savarese, Philipp Heher, Elise N. Engquist, Liam McGuire, Mridul Johari, Gian F. DeNicola, Anne Bigot, Vincent Mouly, Sara Bortolani, Eleonora Torchia, Mauro Monforte, Domenica Megalizzi, Andrea Sabino, Enzo Ricci, Emiliano Giardina, Peter S. Zammit, Giorgio Tasca

**Author notes:** Joint First Authorship (Massimo Ganassi, Claudia Strafella). Joint Senior Authorship (Emiliano Giardina, Peter S. Zammit, Giorgio Tasca).

## Abstract

Inherited myopathies are genetic disorders characterised by declining motor function due to progressive muscle weakening and wasting. Recently, pathogenic variants in *PAX7*, the master transcriptional regulator of muscle stem cells, have been associated with myopathies of variable severity, arguing for impaired satellite cell function as the main pathogenic driver. Here, we report the characterisation of two missense *PAX7* variants in a patient with asymmetric, progressive muscle weakness affecting facial, upper and lower body muscles, and myopathic changes on muscle pathology. Despite this disorder closely phenocopied the clinical presentation of Facioscapulohumeral muscular dystrophy (FSHD), genetic and epigenetic profiling was inconclusive for FSHD, and exome sequencing revealed two heterozygous variants in *PAX7*: c.335C>T, (p.Pro112Leu) and c.1328G>A (p.Cys443Tyr). Modelling these *PAX7* variants in human myoblasts resembled the transcriptomic findings found in the muscle biopsy from the patient. Specifically, these *PAX7* variants caused upregulation of splicing factors, increase of mitochondrial reactive oxygen species levels and reduced cell proliferation, arguing for a pathomechanism where diminished satellite cell function impairs muscle homeostasis. Together, multimodal investigation suggests that these variants in *PAX7* are likely causative of an FSHD- like autosomal recessive myopathy and expand the spectrum of neuromuscular disorders originating form impaired satellite cell function.

## Introduction

Skeletal muscles constitute approximately one-third of total body weight in humans and serve multiple functions: sustain the skeletal system, generate force for movement, and support thermoregulation and metabolism[1]. Hence, diseases affecting muscle structure and function significantly reduce quality of life. Due to persistent exposure to mechanical stress, muscle fibres rely on their ability to be locally repaired. Muscle stem cells, named satellite cells, reside dormant adjacent to each muscle fibre and ensure their homeostasis, promptly responding to demand for growth, regeneration and repair. Upon stimulus, satellite cells activate from quiescence, undergo extensive proliferation to generate a new population of myoblasts, which then either differentiate and fuse into existing fibres or together to form new myofibres[2]. A fraction of myoblasts self-renews to maintain the stem cell pool and regenerative potential. Satellite cells are essential to ensure life-long muscle plasticity[3].

Muscular dystrophies and inherited myopathies comprise a heterogeneous group of genetic conditions presenting with progressive muscle weakness and wasting[4, 5]. Muscular dystrophies often exhibit chronic cycles of muscle fibre degeneration followed by inefficient satellite cell-mediated repair, resulting in continued replacement of muscle tissue with fat, immune cell infiltrates and fibrotic tissue[6, 7]. Diminished muscle plasticity is generally common across muscular dystrophies and myopathies indicating progressive loss of satellite cell function. We recently introduced the concept of satellite cell-opathies, for conditions where the pathogenic variants directly impair the function of genes involved in satellite cell-driven muscle regeneration[8, 9]. Myopathic conditions associated with loss-of function variants in the *PAX7* gene, a key regulator of satellite cells transcriptome, are archetypal satellite cell-opathies[10, 11], presenting limited regeneration capacity leading to atrophic myofibres and fibroadipose tissue replacement.

Satellite cell dysfunction may also arise from other detrimental genetic changes affecting both satellite cells and muscle fibres, such as in Facioscapulohumeral muscular dystrophy (FSHD), the third most common muscular dystrophy. FSHD usually presents with slowly progressing, asymmetric muscle weakness and wasting affecting facial, shoulder girdle and proximal upper limb muscles next spreading to lower limbs[12–14]. Disease inheritance is mainly autosomal dominant and associates with epigenetic derepression at the subtelomeric region of chromosome 4 (4q35), covering a large macrosatellite array of *D4Z4* repeated units (RU) that are normally transcriptionally silenced through DNA methylation[15–17]. FSHD is subclassified into FSHD1 (OMIM: 158900), representing 95% of cases, with the 4q35 locus bearing only 1-10 *D4Z4* units from the ≥11-100+ copies found in non- affected individuals [17, 18], and FSHD2 (OMIM: 158901), displaying a number of *D4Z4* RU usually within the lower end of the normal range and epigenetic derepression primarily caused by concomitant disruptive variants in the chromatin remodelling gene *SMCHD1*, and more rarely in *DNMT3B* or *LRIF1*[19–22]. Each 3.3 kb *D4Z4* RU contains an open reading frame encoding a transcription factor called *Double Homeobox 4 (DUX4*). Demethylation permits expression of the *DUX4* retrogene from the most distal *D4Z4* unit, with mRNA stabilisation by a polyadenylation signal located in the flanking DNA of permissive 4qA haplotypes[23, 24]. Thus, mis-expression of *DUX4*, and its target gene-signature, which is more consistently found in muscles showing signs of disease activity identified on short- tau inversion recovery (STIR) on muscle magnetic resonance imaging (MRI)[25, 26], strongly associate with FSHD pathogenesis[27–30]. In-depth transcriptomic analysis also revealed a concomitant repression of PAX7-target genes that efficiently correlates with disease progression[31–33], implying that FSHD pathogenesis may also involve a satellite cell component possibly arising from DUX4/PAX7 mutual inhibition converging on muscle repair mechanisms[34]. FSHD muscles can display features of impaired regeneration[35, 36], further suggesting that altered satellite cell function or PAX7 activity may result in muscle alterations akin to FSHD.

Here we describe a myopathy with progressive asymmetric muscle involvement, phenocopying FSHD. Genetic testing for FSHD using Southern blotting and optical genome mapping did not identify any disease-related genetic signature and were overall inconclusive for FSHD, suggesting that the FSHD-like pattern could arise from a DUX4-independent mechanism. Histological analysis showed mild myopathic changes and almost complete absence of regenerative fibres in patient’s biopsies. Whole exome sequencing identified compound heterozygous missense variants at highly conserved residues in important domains in PAX7 as the most likely pathogenic finding. Modelling these variants showed changes in cellular function and reduced myoblast proliferation, pointing to satellite cell dysfunction as the cause of impaired muscle regeneration.

## Results

### Clinical presentation

The proband was a male born at term from non-consanguineous parents. The delivery was uncomplicated and there was no history of congenital weakness or other medical problems at birth and in the first years of life. Disease onset was during mid adolescence, when he noticed reduced muscle mass of the right biceps brachii and weakness in elbow flexion that later progressed to shoulder girdle muscles with difficulties in raising the right arm. He reported wasting and weakness in the right quadriceps since later teenage years, as well as difficulties in finger extension of the right hand at older age (mid 20’s). Medical history was otherwise unremarkable, and family history was negative for neuromuscular diseases. Serum creatine kinase level (CK) was 1-1.5 normal values and electromyography was myopathic.

The proband has been followed since his late teen years, and on the last clinical examination (late 30’s), he showed difficulties in walking on heels on the right, diffuse hypotrophy of scapular girdle, both pectoralis, right quadriceps and gastrocnemius muscles, and scapular winging. He could abduct his arms up to 80 degrees. He showed weakness of facial muscles (orbicularis oris), as well as of biceps brachii (Medical Research Council, MRC, 2 on the right and 3 on the left), and clearly asymmetric weakness of finger and wrist extensors (MRC 3), intrinsic hand muscles (MRC 4), knee flexors (MRC 2), tibialis anterior (MRC 2), and extensor hallucis longus (MRC 3), all on the right side (Figure 1A). No tendon contractures or scoliosis were present. Muscle MRI of upper [37] and lower body [38] at proband’s mid 30’s confirmed the prominent asymmetry and showed a pattern highly suggestive of FSHD, with trapezius, serratus anterior, and pectoralis muscles involvement (Figure 1B,C). In addition, upper body imaging highlighted the sparing of subscapularis, supra- and infraspinatus muscles, all features significantly associated with FSHD[37] (pattern 1 in [39]) (Figure 1B,C). In the lower body scan, there was prominent wasting of abdominal musculature (Figure 1D), posterior thigh and anterior leg on the right side (Figure 1E-F), with right tibialis anterior presenting hyperintense signal on STIR sequences indicative of muscle oedema (Figure 1G). Minor signal changes and hypotrophy compared to the contralateral were found in the iliopsoas muscle, which was otherwise largely preserved (Figure 1D). In absence of these latter abnormalities, the combination of involvement would have been pathognomonic for FSHD (pattern 3 in [39]).

**Figure 1.**
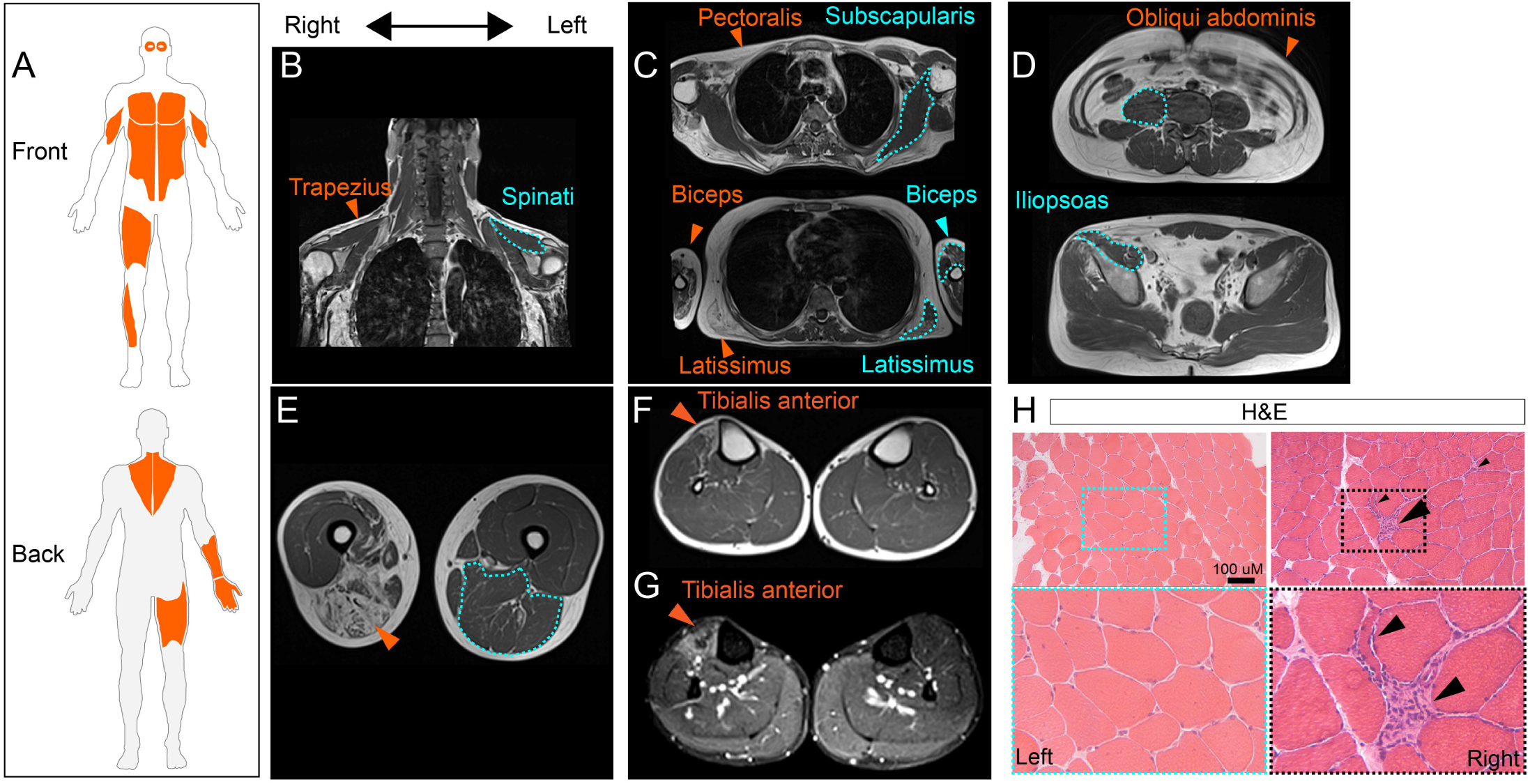
Clinical presentation, muscle imaging and histopathology. **A.** Schematic of affected front/back musculature in the proband suggestive of an FSHD pattern (areas highlighted in orange). **B-G.** Muscle MRI scan in proband mid-30’s. Right/left orientation is reported in coronal (B) and axial (C-G) scans E. T1-weighted (B-F) and STIR (G) images. Acquisitions in the upper body showed trapezius (B, arrowhead) and pectoralis wasting (C, arrowhead) with sparing of spinati (B, dashed line) and subscapularis (C, dashed line) muscles. Biceps brachii (C, arrowhead) and latissimus dorsi (C, arrow) were also asymmetrically involved (arrowheads indicate severely involved muscles on the right and dashed lines less affected muscles on the left). In the lower body, selective involvement of obliqui abdominis (D) was present on both sides, while iliopsoas was largely preserved, with only minor changes on the right (D, dashed line). The right thigh was significantly hypotrophic compared to the left one and displayed severe fatty replacement of adductors and posterior compartment muscles (E, arrowhead). Finally, the asymmetric involvement of the right tibialis anterior (F, arrowhead), which also presented hyperintense signal on STIR sequences (G, arrowhead), was the main abnormality in the lower leg. **H**. Representative haematoxylin and eosin (H&E) staining highlighting the presence of necrotic fibres invaded by macrophages (arrowhead), scattered angulated fibres (small arrowheads), and overall increase in fibre size variability in the biopsy from right thigh muscle compared to the left.

Given the striking side-to-side asymmetry, two biopsies were performed on the right and left vastus lateralis to compare affected and less affected sides and to further investigate possible variants of interest at the transcriptomic level. Histological analysis of muscle biopsies from both vastus lateralis muscles showed myopathic changes (increase in fibre size variability, hypotrophic fibres, mild increase in myofibres with internal nuclei) which were more prominent on the right side. The latter sample also showed scattered necrotic fibres invaded by macrophages (Figure 1H).

### Genetic testing identifies *PAX7* compound heterozygous variants

Based on the clinical and radiological suspicion of FSHD, genetic testing for FSHD1 and 2 was carried out on the affected proband. The EcoRI fragments were 120 kb (33 *D4Z4* Repeat Units; RU) and 85 kb (23 *D4Z4* RU) in length, both with a 4qA haplotype (Supplementary Figure 1A, B). Optical Genome Mapping was also performed on the proband, confirming both the haplotype and sizing of the *D4Z4* alleles, as well as exclusion of more complex rearrangements associated with FSHD, such as in-*cis D4Z4* duplications, mosaicisms undetectable by Southern blotting or copy number variations in proximity of the *SMCHD1* gene on chromosome 18. (Supplementary Figure 1A) In addition, we assessed methylation profile at *D4Z4* locus and found features that were non-conclusive for either FSHD1 or 2[40] (Supplementary Figure 1C). Overall, genetic and epigenetic testing did not confirm a diagnosis of FSHD.

We next carried out whole exome sequencing (WES) to investigate pathogenic and likely pathogenic variants in a panel of genes (>1400) previously associated to other myopathies or candidate disease modifiers for FSHD (Supplementary Table 1). WES identified two heterozygous variants in *PAX7* (NM_001135254.2): c.335C>T, p.Pro112Leu (P112L) and c.1328G>A, p.Cys443Tyr (C443Y) (Figure 2A).

**Figure 2.**
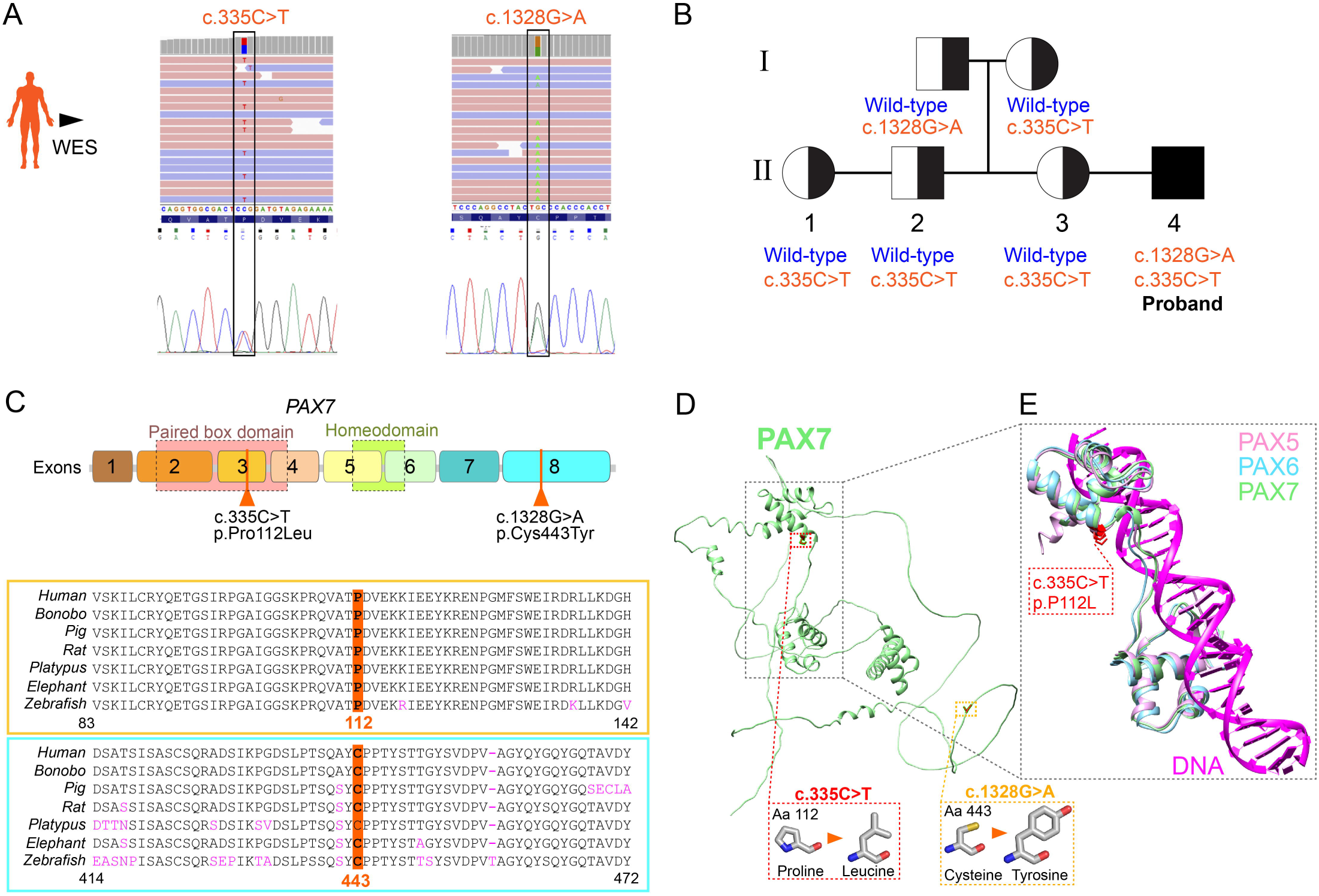
Genomic analysis reveals compound heterozygous alleles in PAX7. **A.** Identification of PAX7 variants, c.335>T (exon 3) and c.1328G>A (exon 8) through whole exome sequencing (WES) and confirmed by sanger sequencing with chromatograms showing the variants alongside their corresponding nucleotide coding positions are presented. **B.** Family pedigree showing *PAX7* alleles (wild-type or variants) in proband family members. The affected individual (proband, filled symbol) carried both variants in *PAX7* in compound heterozygosity. Individuals I:2, II:1,2,3 carried the heterozygous c.335C>T variant only, while I:1 is heterozygous for the c.1328G>A variant. **C.** Schematic of PAX7 coding sequence with numbered exons, domains and positions of the c.335C>T (exon 3) and c.1238G>A variants. Amino acid changes are reported. Below, alignment of PAX7 paralog sequences across indicated species shows high conservation of Proline 112 and Cysteine 443. Pink letters highlight residue changes relative to human PAX7 sequence. Below are diagrams of amino acid residues to highlight structural changes. **D.** Ribbon representation of the human PAX7 protein AlphaFold model. The position of the variants on the three-dimensional structure is highlighted. Cysteine 443 is located within the C-terminal region of PAX7 which is predicted to be unstructured. **E.** Superimposition of the AlphaFold structure of conserved Paired domains and proximal regions of PAX7 and its orthologs PAX5 and PAX6 in complex with DNA (PDBID: 6pax, 1mdm). PAX7 is in green, PAX5 in purple, PAX6 in cyan and the DNA in magenta. The site of the P112L mutation is shown in red. Proline is conserved in PAX5,6,7 and in close vicinity of protein loop binding to DNA.

Segregation analysis of the variants in healthy family members revealed that the variants co-segregated with the disease: all siblings (II:1, II:2, and II:3) and the mother (I:2) were heterozygous for the c.335C>T variant, while only the father (I:1) carried the c.1328G>A variant, therefore supporting a likely autosomal recessive mode of inheritance, since single allele heterozygosity for either variants alone is non-pathogenic (Figure 2B). Analysis of the variants using Exomiser[41] and HPO terms related to myopathy also prioritised both the gene and its variants as clinically relevant.

The c.335C>T (P112L) variant resides within the Paired box domain, responsible for DNA binding and motif recognition on specific PAX7 target genes, whereas c.1328G>A (C443Y) locates within exon 8 (Figure 2C). Both residues are highly conserved in PAX7 paralogs across several vertebrate species indicating their importance. The c.335C>T variant has a Minor Allele Frequency (MAF) of 0.002 in gnomAD/ExAC/1000Genomes and is predicted to be deleterious by REVEL[42]. The Proline-to-Leucine substitution (from a rigid side chain important for protein conformation to an aliphatic, hydrophobic side chain that tends to localise within the protein’s internal structure, Figure 2D) is highly unfavored in terms of conserved aminoacidic properties.The c.1328G>A variant is absent in gnomAD, ExAC and1000 Genome databases as well as in gene variant databases (ClinVar and LOVD) It is predicted to be deleterious and to affect protein function due to the significant aminoacidic substitution from Cysteine (side chain able to form disulphide bond critical for protein stability maintenance) to Tyrosine (aromatic side chain which can modify protein interactions).

Structural modelling highlighted that Proline 112 is conserved across PAX7 orthologs PAX5 and PAX6 and locates proximally to the DNA-binding loop, suggesting that the P112L variant could affect the positioning of the loop interacting with DNA, thereby impairing PAX7 transcriptional activity at target genes (Figure 2D,E). Tyrosine is significantly larger than Cysteine implying that C443Y could affect PAX7 folding, impairing protein function. However, Cysteine 443 is in an unstructured region (ensembl.org) so the effect of this substitution remains unclear (Figure 2D).

### The *PAX7* variants do not reduce satellite cell number

Since disorders related to loss of PAX7 present altered proportions of satellite cells [10, 11], we tested this hypothesis on both biopsies comparing them with a normal (healthy) control, an immune-mediated muscle disorder (Necrotising Autoimmune Myopathy: NAM), and two FSHD samples showing either active (STIR^+ve^) and non-active (STIR^-ve^) disease.

Immunolabelling for PAX7 indicated a number of satellite cells comparable with that found in the healthy control sample, NAM and the FSHD STIR^-ve^. In contrast, the FSHD STIR^+ve^ specimen displayed a higher number PAX7^+ve^ satellite cells, suggestive of an active regeneration process (Figure 3A). These results indicate that PAX7 variants do not significantly alter the number of PAX7^+ve^ satellite cells.

**Figure 3.**
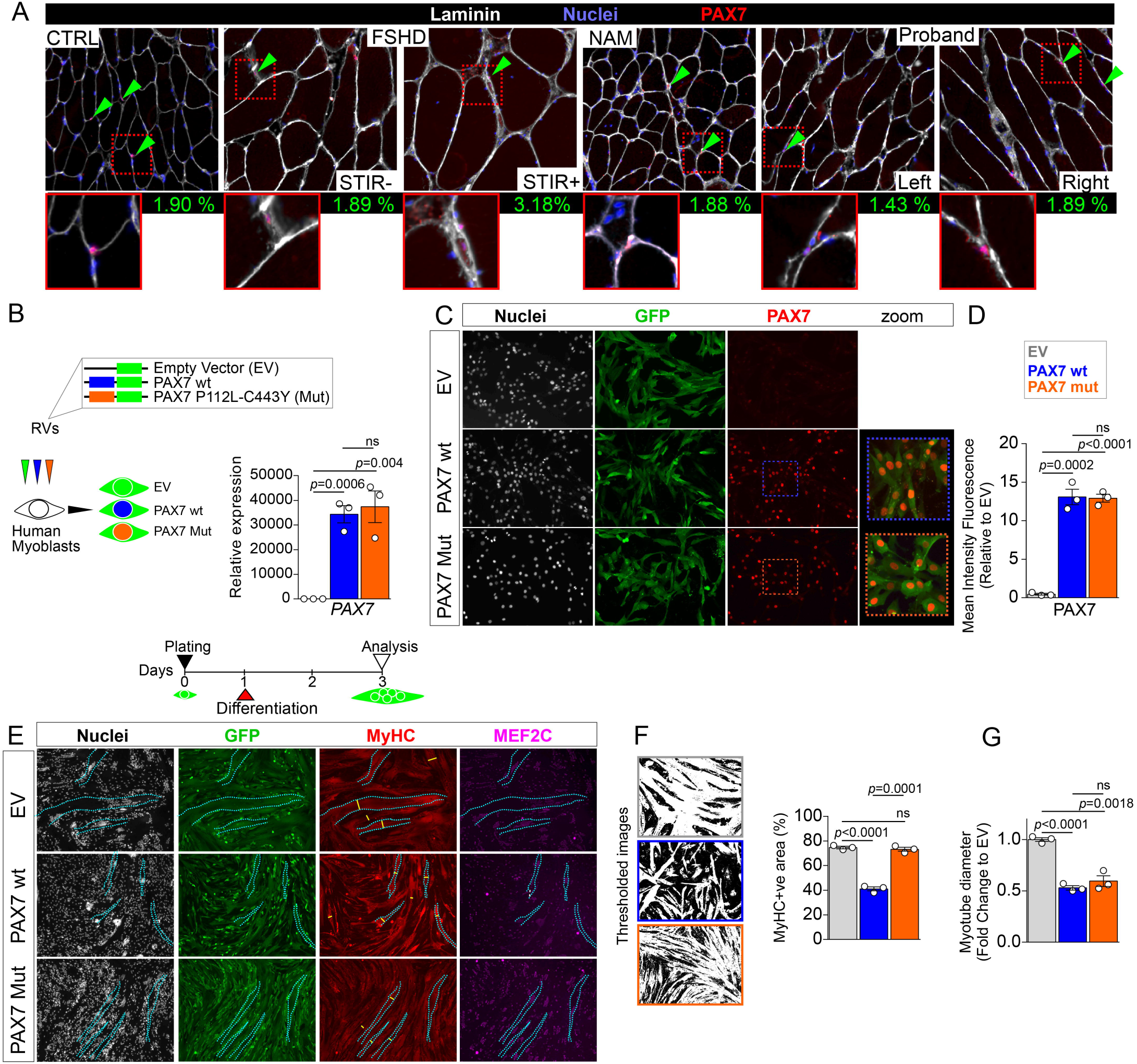
P112L-C443Y variant alters PAX7 function without altering its stability. **A.** Representative images of immunolabelling for PAX7 (red) counterstained with Wheat Germ Agglutinin (WGA, white) and Hoechst (blue) showing satellite cells, fibre boundaries and nuclei respectively in different biopsies (CTRL: healthy; NAM: Necrotising Autoimmune Myopathy; FSHD STIR^+ve^: active; and FSHD STIR^-ve^: non- active disease). Percentage of PAX7^+ve^ nuclei (satellite cells) over the total nuclei in the section is reported per each biopsy. Green arrowheads indicate satellite cells, with representative images shown in insets. **B.** Schematic of retroviral infection to obtain stable cell lines, encoding for either PAX7 wild-type (PAX7wt) or PAX7 P112L-C443Y (PAX7mut), and GFP alone (empty vector; EV). RT-qPCR confirming increased PAX7 expression in both PAX7wt and PAX7 mut cell lines compared to empty vector control cells. **C.** Representative images of proliferating EV, PAX7wt or PAX7mut myoblast lines immunolabelled for GFP (green), PAX7 (red) and nuclei counterstained with Hoechst (white), showing clear accumulation of PAX7 in PAX7wt or PAX7mut cells. **D.** Quantification of PAX7 nuclear staining confirms similar accumulation and nuclear localisation in both PAX7wt or PAX7mut cell lines. **E.** Schematic of experimental design for differentiation analysis. Myoblasts were plated at equal numbers and induced to differentiate for 2 days, prior to immunolabelling. Representative images of myotubes from EV, PAX7wt or PAX7mut differentiated myoblasts immunolabelled for MyHC (red), GFP (green), MEF2C (magenta) and nuclei counterstained with Hoechst (white), showing severely reduced myotube formation in PAX7wt compared to PAX7mut and EV control cells. **F.** Example of thresholded images from E used for quantification of MyHC labelling. MyHC^+ve^ area is dramatically reduced in PAX7wt differentiated cells, but not in PAX7mut compared to EV control. **G.** PAX7wt or PAX7mut myoblasts differentiate in myotube with significantly reduced diameter myotubes compared to EV control. All graphs report unpaired t-test analysis.

### P112L-C443Y PAX7 is normally expressed and localised in myoblasts

Next, we assessed the effect of the detected *PAX7* variants on myogenesis. Since PAX7 binds to DNA as cooperative dimers[43–46] and considering that variants did not cause disease in heterozygous carriers, human myoblasts were transduced with retrovirus encoding either *PAX7* bearing both variants found in the proband (PAX7mut) or wild-type *PAX7* (PAX7wt) as control, and stably expressing lines were made (Figure 3B). RT-qPCR analysis confirmed significant upregulation of *PAX7* mRNA in both PAX7wt and PAX7mut lines, compared to control myoblasts transduced with the empty vector (EV). Since both PAX7wt and PAX7mut were expressed at comparably high levels compared to EV, we concluded that the variants do not affect the stability of *PAX7* mRNA (Figure 3B). Immunolabelling PAX7 also confirmed significant accumulation of PAX7 protein in nuclei of PAX7wt and PAX7mut myoblasts (Figure 3C,D). Thus, the combination of P112L-C443Y variants does not alter expression or nuclear localisation of PAX7 in human myoblasts.

### P112L-C443Y variants reduce PAX7 inhibitory function on myogenic differentiation

Structural modelling suggested that P112L-C443Y variants alter PAX7 function. Since PAX7 inhibition on myogenic progression is well established[47–49], we assessed the effect of P112L-C443Y on differentiation of human myoblasts by immunolabelling for Myosin Heavy Chain (MyHC), a marker of terminal differentiation. Human myoblasts differentiated for two days showed a significant reduction in myotube formation and growth in PAX7wt compared to EV controls (Figure 3E-G), confirming previous findings in murine cells[50]. Levels of MEF2C, a key transcription factor in myogenesis[51–53], were also reduced, further confirming PAX7-driven repression of the differentiation program in human myogenesis. In contrast, PAX7mut myoblasts displayed greater differentiation capacity, as measured by MyHC area coverage, suggesting a loss-of PAX7 inhibitory function compared to wt control. However, the diameter of myotubes in PAX7mut was comparable to that of PAX7wt, which were both reduced compared to EV controls (Figure 3G), suggesting that PAX7mut maintains an inhibitory function on myotube growth. We concluded that the P112L-C443Y PAX7 variant alter protein function(s) likely resulting in impaired myogenesis.

### P112L-C443Y PAX7 expressing human myoblasts recapitulate key transcriptomic abnormalities found in proband’s muscle

To gain further insight into the pathomechanism of myopathy, we deployed transcriptomic analysis of the two different muscle biopsies from the proband and compared them to three unrelated individuals with no muscle pathology. The sample from the right quadriceps was split and ran in two separate experimental batches. Principal component analysis on the gene expression data confirmed efficient separation of proband samples compared to controls, with the two biopsies of the right muscle clustering together, slightly separated from the one from left muscle (Figure 4A). Transcriptomic analysis retrieved 885 differentially expressed genes in the proband biopsies (476 Up and 409 Down). Given the similarity to an FSHD clinical phenotype, we analysed transcriptomic data of the proband for the presence of the DUX4 signature, ie. DUX4-target genes found upregulated in FSHD muscles[54]. Only two DUX4 target genes, namely *TENT5C* and *CXCR4,* were differentially regulated in the proband although in opposite direction compared to that expected with DUX4 activation (Figure 4B), and *DUX4* itself was undetectable (not shown).

**Figure 4.**
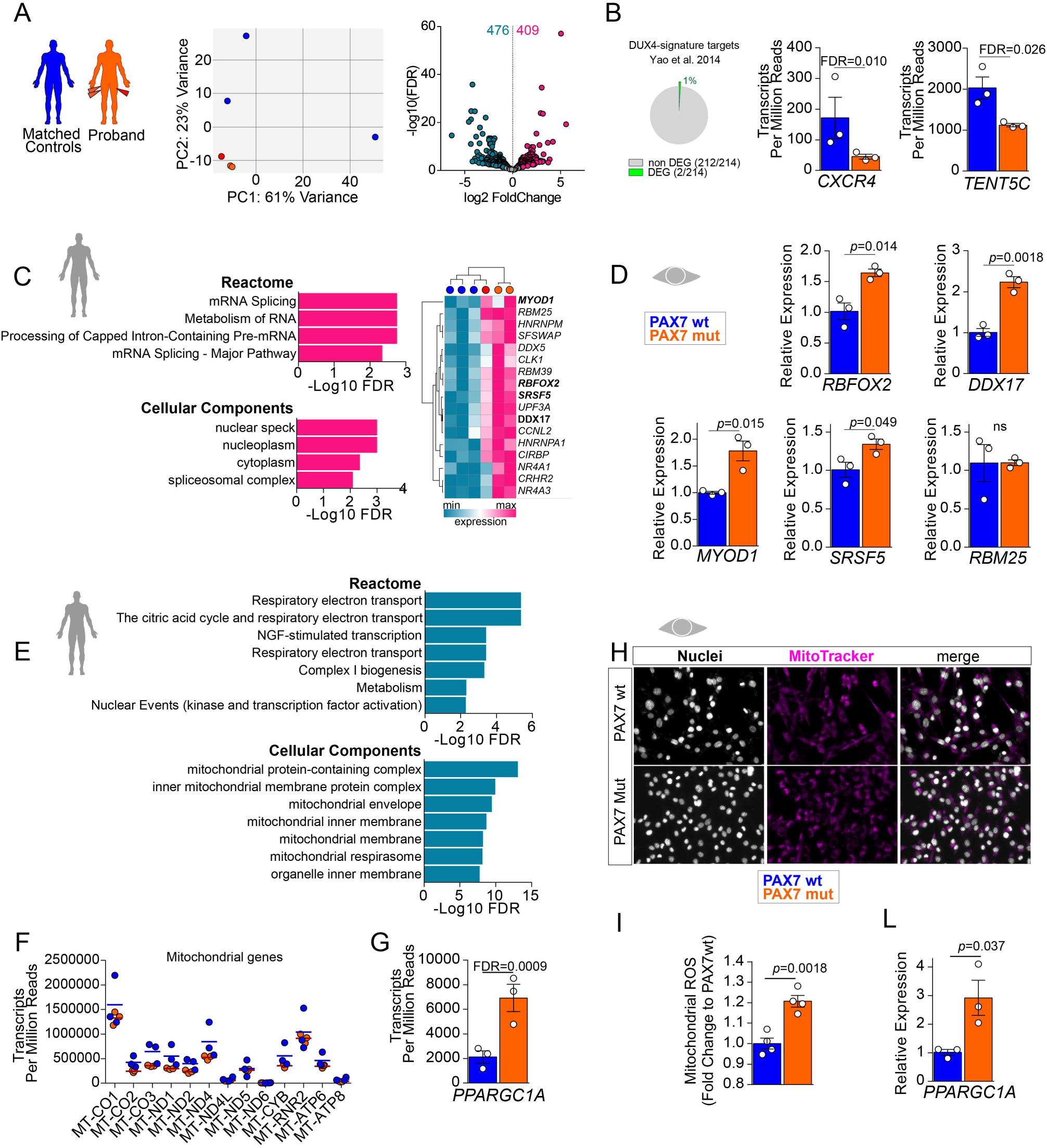
Expression of PAX7 P112L-C443Y in myoblasts reproduces key transcriptomic features found in the proband. **A.** Schematic of experimental setup and PCA analysis on transcriptomes from patient and control samples. Volcano plot reports significant DEGs (differentially expressed genes) with *p*[≤[0.05 and log2FC[≥[0.5 between patient’s biopsies and three control samples. The number of up- (pink) and down- regulated (light blue) DEGs is reported. **B.** Comparative transcriptomic analysis showing 2 differentially expressed genes out of the 213 constituting the DUX4-target gene signature from[54] in the proband’s transcriptome. Expression values (Transcripts Per Million reads) of *CXCR4* and *TENT5C* in patient transcriptome compared to control samples are reported. **C.** Gene Ontology analysis on up-regulated genes (pink in A) in proband muscle biopsy samples showing annotation in indicated datasets. Expression of the unique 17 genes listed in *Reactome* GOs referring to mRNA splicing in proband (orange) or control muscle (blue) biopsies efficiently separated by hierarchical clustering analysis. **D.** RT-qPCR showing significantly increased expression of indicated genes in PAX7mut myoblasts compared to PAX7wt. **E.** Gene Ontology analysis on down-regulated genes (light blue in A) in proband muscle biopsy samples showing annotation in indicated datasets. **F.** Expression of 13 protein-encoding mitochondrial genes in the muscle biopsies from proband (orange) and three independent control muscle samples (blue). **G.** Transcriptomic analysis showing significantly increased *PPARGC1A* (*PGC1A*) in proband biopsies. **H.** Representative mitochondrial staining in PAX7wt and PAX7mut cells indicate no overt defects in mitochondria. **I.** Quantification of Mitochondrial Reactive Oxygen Species (mitoROS) shows a significant increase in PAX7mut compared to PAX7wt control cells. **L**. RT-qPCR showing an increased *PGC1A* expression in PAX7mut myoblast cell line compared to PAX7wt. All graphs report False discovery rate (FDR, q-value) or unpaired t-test analysis (p- value).

Next, we interrogated the 476 Up and 409 Down regulated genes in proband biopsies using Gene Ontology (GO) analysis (Supplementary Table 2). The list of upregulated genes converged on mRNA regulation and splicing, with several splicing factors being upregulated in the proband compared to healthy controls (Figure 4C). We then performed RT-qPCR on our cellular model and found the expression of *MYOD1*, *RBFOX2*, *DDX17* and *SRSF5* upregulated in PAX7mut myoblasts (Figure 4D).

Analysis on downregulated genes retrieved GOs related mainly to mitochondrial function, such as ‘*Respiratory electron transport*’, ‘*The citric acid cycle*’ and ‘*Metabolism*’ arguing for a contribution of mitochondrial impairment to muscle pathology (Figure 4E). The expression pattern of the 13 protein coding genes in the mitochondrial genome involved in the respiratory chain composition was comparable to control sample biopsies (Figure 4F), suggesting that potential direct transcriptional effects on mitochondrial respiratory chain function were driven by altered expression of nuclear genome genes. Indeed, the expression of *PPARGC1A*, encoding for the master regulator of mitochondrial homeostasis PGC1A, was significantly upregulated in proband biopsies (Figure 4G), arguing for abnormal mitochondrial homeostasis. Coherently, while PAX7mut myoblasts did not show an overt mitochondrial phenotype compared to PAX7wt (Figure 4H), mitochondrial Reactive Oxygen Species (mitoROS) levels were significantly increased in PAX7mut cells (Figure 4I), and paralleled by elevated *PGC1A* expression (Figure 4L), resembling transcriptomic features observed in proband muscles. Overall, these results provide further support that the molecular alterations observed in the proband may rely on a PAX7-driven pathomechanism likely altering satellite cell activity.

### P112L-C443Y PAX7 alters viability of human muscle cells

Since PAX7 is a master regulator of satellite cell function, we reasoned that P112L- C443Y-induced cellular alterations may impair myoblast viability. We set out to compare proliferation rate of PAX7wt and PAX7mut myoblasts by plating at equal numbers and culturing in growth medium for three days. Longitudinal analysis revealed that both cell lines significantly increased in number over time (Figure 5A), indicating that PAX7mut variant does not induce cell toxicity. However, comparison of proliferation rate highlighted that PAX7mut myoblasts proliferated less compared to PAX7wt control cells (Figure 5B). To explore proliferation dynamics upon PAX7 P112L-C443Y expression, PAX7wt and PAX7mut myoblasts were pulsed with EdU for 2 h after three days in growth medium, and EdU incorporation was compared (Figure 5C). The proportion of cells in S-phase was significantly decreased in PAX7mut compared to PAX7wt (Figure 5D), indicating that PAX7 P112L-C443Y impairs cell cycle progression. Parallel RT-qPCR analysis highlighted higher expression of the negative regulator of the cell cycle *RB1* (Figure 5E), arguing for P112L-C443Y-induced cell-cycle alteration through transcriptional regulation.

**Figure 5.**
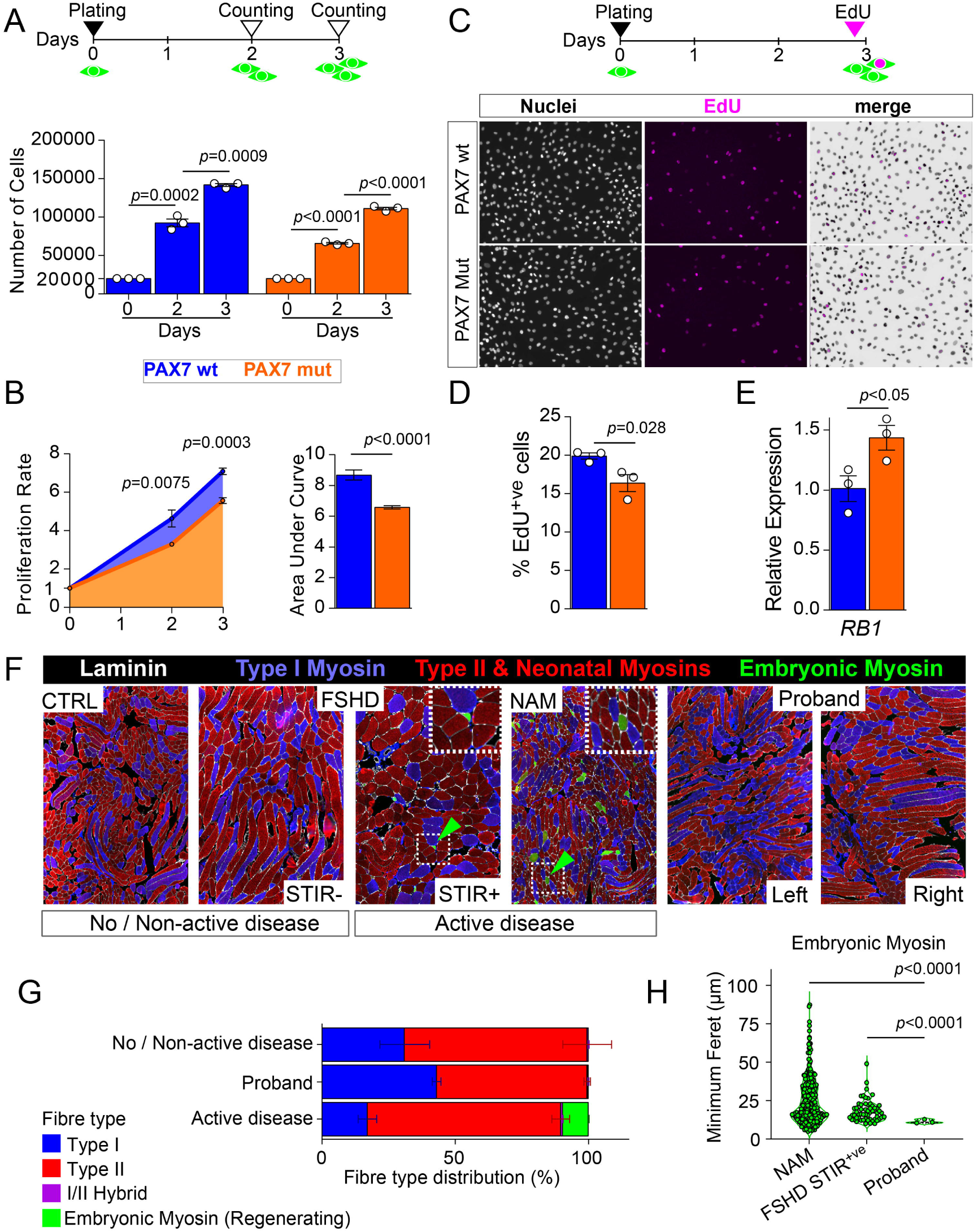
PAX7 P112L-C443Y decreases myoblast proliferation and could contribute to reduced muscle regenerative potential in the proband. **A.** Schematic of experimental design for viability analysis. Myoblasts were plated at equal numbers and allowed to expand in proliferation medium. Number of cells was counted at 2 or 3 days after plating (day 0). (Bottom) Quantification of number of cells indicates that all three myoblast lines (Empty vector (EV), PAX7 wild type (wt), PAX7 P112L-C443Y (mut)) are viable and proliferate in culture over time. **B.** Quantification of cell proliferation from day 0 to day 3 from A (left) showing reduced rate in PAX7mut myoblasts compared to PAX7wt. Quantification of areas under curve (coloured area underneath blue (PAX7wt), orange (PAX7mut)) shows overall proliferation dynamics indicates reduction in PAX7mut myoblasts. **C.** Schematic of experimental design for proliferation analysis. Cells were plated and cultured as in A, and EdU pulsed at day 3 for 2 hours before being fixed for analysis. Representative images of proliferating PAX7wt and PAX7mut myoblasts stained for EdU (magenta) and nuclei counterstained with Hoechst (white), showing reduced EdU accumulation in PAX7mut myoblasts. **D.** Quantification of fraction of EdU^+ve^ myoblasts after 3 days in proliferating condition confirms significantly reduced proliferation rate in PAX7mut compared to PAX7wt. **E.** RT-qPCR showing significantly increased *RB1* expression in PAX7mut myoblast cell line compared to PAX7wt. **F.** Representative images of immunolabelling for Slow (blue), Fast and Neonatal (red), and embryonic myosin heavy chain (MYH3), together with Laminin (white), to highlight fibre types and fibres boundaries in the different biopsies examined. **G.** Comparison of fibre type composition across ‘No/Non-active disease’ (CTRL, FSHD-STIR^-ve^), ‘Active disease’ (FSHD-STIR^+ve^, NAM) and proband identified regenerating fibres mainly in the ‘Active disease’ group. **H.** Quantification of regenerating myofibre (MYH3^+ve^) diameter (minimum Feret) highlighted reduced size in proband biopsies compared to FSHD-STIR^+ve^ and NAM. Regenerating fibres were absent in control and FSHD-STIR^-ve^ samples. All graphs report unpaired t-test analysis.

We reasoned that impaired satellite cell proliferation would hamper regeneration capacity and contribute to the progressive myopathy observed in the proband. Hence, we assessed fibre-type composition in our cohort of muscle biopsies, pooling samples in our panel for ‘*No or non-active disease*’ (CTRL and FSHD-STIR^-ve^), *active disease*’ (FSHD-STIR^+ve^ and NAM), and compared those with proband biopsies. Fibre-type composition was similar across samples (Figure 5G), with FSHD-STIR^+ve^ displaying an increased fibre size compared to the other samples. In contrast, regenerating fibres (embryonic eMyHC^+ve^) were found almost exclusively in the ‘*active disease*’ samples (Figure 5F,G). The few regenerating myofibres identified in the proband muscles displayed significantly reduced size compared to those found in NAM and FSHD-STIR^+ve^, arguing for reduced or inefficient muscle regeneration (Figure 5G). No regenerating fibres were present in CTRL and FSHD-STIR^-ve^ samples.

Notably, transcriptomic analysis retrieved no changes in expression of *PGDFRa*, *PAX3* and *CLEC14A* (Supplementary Figure 2), previously found upregulated in patients bearing pathogenic *PAX7* null mutations[10, 11], excluding activation of compensatory pathways for muscle regeneration. Likewise, expression of *PAX7* was comparable between proband and controls (Supplementary Figure 2), in line with the unchanged number of satellite cells in muscle biopsies and confirming that the combination of *PAX7* P112L and C443Y variants do not affect gene expression or RNA stability as observed for other mutations previously reported[10].

Our results suggest that PAX7 P112L-C443Y compound heterozygous variants can impact muscle proliferation, leading to inefficient muscle regeneration and progressive muscle weakening and wasting.

## Discussion

Here we report that compound heterozygous *PAX7* variants (P112L and C443Y) associate with a myopathic phenotype akin to FSHD characterized by progressive muscle wasting, declining muscle function and possibly impaired muscle regeneration. These findings support the inclusion of PAX7-RD (Related Diseases) to the list of differential diagnoses of an FSHD-like phenotype and negative FSHD1 and 2 genetic testing.

The clinical assessment of the proband highlighted a pattern of muscle involvement that resembled typical FSHD [37, 38] with asymmetric, upper limb predominant wasting associated with scapular winging and facial muscle weakness with onset in the second decade. However, extensive molecular testing failed to confirm an FSHD diagnosis, and in-depth transcriptomic analysis also did not reveal any specific abnormality potentially attributable to DUX4 upregulation [54].

Interestingly, we identified biallelic variants in *PAX7* as the possible cause of the muscle disease in the proband. Segregation analysis confirmed the cosegregation of the variants with the disease in the family, proving also that carriers of each variant do not show any phenotype.

Different muscular and non-muscular phenotypes have been described so far as associated with pathogenic *PAX7* variants (Supplementary Table 3). Homozygous variants within exons 2 and 3, encoding for the Paired domain (R74*, R56C and R145*), have been linked with MYOSCO (CMYO9, OMIM: 618578), a satellite cell- opathy presenting with congenital progressive muscle weakness and atrophy with scoliosis and dysmorphic facial features, paralleled by a severe reduction in the satellite cell pool[8–11, 55]. A homozygous missense variant downstream of the homeodomain (N267K) was recently associated with congenital myopathy overlapping with MYOSCO, expanding the spectrum of PAX7-related muscle conditions[56].

Single heterozygous variants in residues following PAX7 homeodomain (A282V and P306L) or surrounding the region of exons 8 and 9 (G459D, Y495T, V454M, G463S) [57], nearby the C443 residue mutated in our proband, have been reported in patients with congenital scoliosis, although no segregation study has been performed to support the pathogenicity of these variants. Primary muscle involvement was also not investigated in detail in these patients showing vertebral malformations.

At variance from these observations, spine involvement was absent either in the proband or in any of the first-degree relatives, indicating that P112L and C443Y *per se* or in compound combination do not directly cause spine deformities or major weakness of relevant axial muscles. Additional heterozygous PAX7 variants in residues surrounding C443 were found in studies investigating orofacial clefting[55, 58–61], with no obvious involvement of skeletal muscles. This is in line with the absence of muscle phenotype in the proband father presenting a heterozygous C443Y allele and argues for significant muscle pathology only arising from either homozygous *PAX7* functional null mutations as seen in MYOSCO or resulting from compound heterozygous effect as in our proband.

Despite progressive exhaustion of the satellite cell pool due to mutations altering PAX7 expression/accumulation, MYOSCO muscles show regenerating (MYH3^+ve^) muscle fibres indicating ongoing, albeit inefficient, myofibre regeneration likely supported by other cell populations (e.g. PDGFRa^+ve^, PAX3^+ve^, CLEC14A^+ve^) compensating for the lack of PAX7^+ve^ cells[10, 11, 62]. Conversely, our proband biopsies showed limited regeneration and no major changes in satellite cell content. Expression of *PDGFRa*, *PAX3*, *CLEC14A* was also consistently unchanged, arguing against compensatory pathways activated upon significant alterations of *PAX7* expression or satellite cell density. We concluded that the variants identified in the proband do not significantly alter PAX*7* mRNA levels, protein content and localization, nuclear localisation, or number of satellite cells. Overall, these pieces of evidence indicate that different alterations of PAX7 structure and function result in different skeletal muscle diseases with peculiar clinical, cellular and molecular features depending on the different functional impacts, expanding the spectrum of PAX7-RD.

Considering the significant phenotypic overlap with FSHD, we investigated whether the partial loss of PAX7 function caused cellular perturbations similar to those driven by DUX4 in FSHD. It has been suggested that both DUX4 and PAX7 may mutually inhibit the activation of their respective transcriptional target genes due to high sequence similarity between the homeodomains [63], and that DUX4 expression in FSHD would hamper PAX7 function leading to poor muscle homeostasis, repair and regeneration accounting for the progressive muscle loss.

Recent evidences coming from studies on patient biopsies^59^ as well as a meta-analysis of several omics datasets of FSHD muscles confirmed that mitochondrial perturbations are present and may contribute to disease pathophysiology[64, 65], and increased production of mitochondrial Reactive Oxygen species as been shown in cultured FSHD myoblasts and upon DUX4 overexpression [66, 67]. We speculate that these mechanisms can constitute a possible molecular link between the consequences of *PAX7* P112L-C443Y variants and mechanisms of muscle damage activated in FSHD, ultimately contributing to the phenotypic overlap.

Another overlapping molecular feature between FSHD and the myopathy in the proband is the alteration of mRNA splicing. The impact of DUX4 on mRNA splicing is considered to be an important contributor to FSHD pathogenesis, with several RNA binding proteins interacting with DUX4 directly[68]. Indeed, misregulation of splicing process is found across several omics datasets arguing for a potentially relevant role of this process in FSHD [64, 65]. Our cellular model argues for *DDX17*, *RBFOX2* and *SRSF5* to respond to accumulation of PAX7 P112L-C443Y variant suggesting a direct activation in satellite cells, which are the only PAX7-expressing/responding cells in skeletal muscle.

DDX17 contributes to mitochondrial homeostasis by controlling ROS production and efficiency of the respiratory chain in different cell types likely via transcriptional regulation[69–71] and was identified as a putative DUX4 interacting partner [72, 73]. RBFOX2 plays a critical role in ensuring mitochondrial health in muscle cells, regulating mRNA levels of mitochondrial factors [74, 75].SRSF5 is a member of serine/arginine-rich protein (SR) family, which can interact with PGC1A[76] and plays key roles in the regulation of pre-mRNA alternative splicing and in cell-cycle progression[77]. Overall, the dysregulation of these three genes may provide a suitable link between PAX7 function and mitochondrial activity converging on cell viability.

Transcriptomic profiling of patient’s biopsies also highlighted concomitant *MYOD1* upregulation. Notably, *MYOD1* regulates skeletal muscle oxidative metabolism and has also a role in governing muscle-specific alternative splicing of the mitochondrial ATP Synthase-Subunit pre-mRNA during myogenesis[78, 79], in line with the mitochondrial dysfunction identified in our study. The increased *MYOD1* level following expression of PAX7 P112L-C443Y further suggests a mechanism through which altered splicing affects mitochondrial function and impair muscle cell proliferation. MYOD1 increase is also likely to be responsible for the upregulation of *RB1* and consequent decrease in cell-cycle progression as reported before in muscle cells[80].

In conclusion, we provide several pieces of evidence indicating that biallelic *PAX7* variants may phenocopy FSHD likely via a novel pathomechanism, implicating satellite cell dysfunction without depletion. Similarly to FSHD, our results are consistent with an interplay between altered expression of splicing factors and mitochondrial dysfunction leading to impaired muscle cell viability in this new disease. Such findings have clinical relevance since they may help with the molecular diagnosis for those patients presenting a classical FSHD involvement pattern lacking the canonical (epi)genetic layout at 4q35, and development of tailored therapies.

## Material and Methods

### Genetics

The genomic DNA of the proband was extracted from peripheral blood sample (35 ml) by manual and automatised techniques in parallel according to the manufacturer’s instructions. Manual extraction allowed obtaining the high-molecular weight DNA embedded into agarose plugs required for *D4Z4* sizing, whereas the automatised method (MagPurix Blood DNA Extraction Kit and MagPurix Automatic Extraction System, Zinexts) was used to extract the DNA for sequencing the FSHD2- associated genes and for methylation analysis. In addition, 1 ml of fresh blood was employed for the optical genome mapping. The genomic DNA of family members was extracted from 400 µl of blood sample by automatised method and employed for segregation analysis and methylation levels assessment.

The *D4Z4* sizing was performed according to the standard procedure [81]. Briefly, the extracted DNA was digested on agarose plugs by restriction enzymes (EcoRI, EcoRI/BlnI and XapI) and separated by PFGE. The *D4Z4* size was measured by Southern Blot and hybridization with p13E-11 probe according to standard procedure ([82]). Optical Genome Mapping was performed by Bionano technology. The Ultra-High Molecular Weight (UHMW) DNA was isolated by a specific extraction system (Bionano sample preparation method) employing a silica-based magnetic disk to attract large DNA molecules. The DNA was then tagged by Direct Labeling Enzyme 1, loaded on a chip and scanned by the Bionano Saphyr instrument. This approach allowed the linearizing and imaging of the DNA molecules that were analysed by a dedicated pipeline (Bionano EnFocus™ FSHD Analysis 1.0) and the genome map generated was compared to the human reference sequence.

Methylation analysis was performed by a specific protocol consisting of Bisulfite Sequencing (BSS) followed by Capillary Electrophoresis (CE) with Amplified Fragment Length Polymorphisms (AFLP) module described[40]. Two regions within the *D4Z4* locus were evaluated for DNA methylation analysis, namely *DUX4*-PAS (located within the most distal part of the array and suggestive of FSHD1) and DR1 (localized 1kb upstream of the *DUX4*-ORF and indicative for FSHD2). In particular, the assay uses the methylation levels related to four CpG sites (*DUX4*-PAS_CpG6, *DUX4*-PAS_CpG3, DR1_CpG1 and DR1_CpG22) to discriminate subjects with reduced methylation levels compatible with FSHD [83].

Whole Exome Sequencing (WES) was performed on the Next-Seq 550 System (Illumina). Library preparation was performed on 30-50ng/μl of DNA using Illumina DNA Prep with Enrichment and Tagmentation according to manufacturer’s instructions. The obtained libraries were sequenced at 2x100 bp and the sequencing quality of the resulting data were expected to reach a Quality score>30 (Q30) for ∼80% of total called bases. For the resulting variants, only those reporting a minimum coverage of 20X were considered eligible for subsequent analysis. The resulting variants were visualised by Integrated Genome Viewer (v.2.18.4) and functionally annotated using BaseSpace Variant Interpreter (Illumina, v. 2.15.0.110) with GRCh37 as genome build reference. Annotated variants were prioritised considering the type (nonsense, missense, frameshift, splicing, loss- or gain-of-function), frequency, localisation in regulatory regions and their pathogenicity scores. To evaluate the rarity of a variant, publicly available reference database (GnomAD/ExAc/1000Genome) were used. In-silico predictive tools (REVEL, Varsite) were utilized to assess the pathogenicity scores. REVEL (Rare Exome Variant Ensemble Learner) is a meta-predictor tool for missense variants consisting of different scores (MutPred, FATHMM v2.3, VEST 3.0, PolyPhen-2, SIFT, PROVEAN, MutationAssessor, MutationTaster, LRT, GERP++, SiPhy, phyloP, phastCons) that are integrated to provide a unique pathogenicity score of the variants of interest [42]. Moreover, Varsite allows predicting the potential impact of missense variants on protein structure and function[84]. The analysis of variants was performed focusing primarily on the variants localized in genes near the *D4Z4* locus, genes being targeted by *DUX4* or functioning as epigenetic regulators of *D4Z4*, as described elsewhere [85]. In addition, WES was analysed to exclude variants in known genes responsible for myopathies and other neuromuscular disorders (Supplementary Table 1).

Finally, confirmation of the *PAX7* variants observed by WES and segregation analysis within the family members were performed by direct sequencing. The DNA was amplified by classical PCR, using the AmpliTaq Gold DNA Polymerase (Applied Biosystems) reagents in a total volume of 25 μL, following the manufacturer’s instructions. Successively, direct sequencing of the amplified samples was performed by BigDye Terminator v3.1 Cycle Sequencing Kit (Thermo Fisher Scientific) and run on ABI3500 genetic analyzer (Applied Biosystems). The results were finally analysed with Sequencing Analysis Software v.7 (Applied Biosystems).

### Transcriptomic analysis on proband biopsies

Library preparations (polyA+ RNA selection using NEBNext Ultra II Directional RNA Library Prep kit E7760 for Illumina) and sequencing were performed at Oxford Genomics Center, University of Oxford. Libraries were multiplexed and sequenced on Illumina NovaSeq 6000 (average 60 M reads, 150 bp paired-end).

Paired-end sequences in FASTQ format were mapped with STAR two-pass method using STAR 2.7.7a (STAR, RRID: SCR_004463) and the index generated from Gencode.v39 human reference and comprehensive gene annotation (primary assembly). Uniquely mapped fragments were quantified by featureCounts (featureCounts, RRID: SCR_012919) using Gencode.v39 primary comprehensive gene annotation. Differential gene expression analysis was performed with DESeq2 (v1.26.0) (DESeq2, RRID: SCR_015687) in Rstudio (v1.2.5019) (RStudio, RRID: SCR_000432) based on R (v3.6.3) (R Project for Statistical Computing, RRID: SCR_001905).

### Retroviral Expression Constructs

Human PAX7 cDNA (PAX7 wt) was cloned into a modified pMSCV-puro vector (Clontech), in which the puromycin resistance gene has been replaced with an internal ribosomal entry site (IRES) preceding the coding sequence for enhanced green fluorescent protein (eGFP) to obtain pMSCV-IRES-eGFP (pMIG) as previously described[29]. Mutations c.335C>T (p.Pro112Leu) and c.1238G>A (p.Cys443Tyr) were introduced on pMIG-PAX7wt-IRES-eGFP by Genewiz Gene Synthesis service (Azenta, UK) to obtain pMIG-PAX7mut-IRES-eGFP bearing both mutations in cis. For production of retroviral particles constructs were transfected in HEK293T cells as described[29].

### Cell Culture

Immortalized human myoblasts (hSkMC-AB1190) [86] were grown in complete proliferation medium: Skeletal Muscle Cell Growth medium (PromoCell) supplemented with 20% heat inactivated foetal bovine serum (FBS; Thermo Scientific), 50 μg/ml Gentamycin (Life Technologies) and 1 unit of the SupplementMix (PromoCell) and passaged at ∼70% confluency to maintain in ‘proliferation’ state. Myoblasts were plated at a density of 5x10^3^ cells/well in flat-bottomed 96 well plates for immunolabelling and at 0.75x10^6^ cells/well in 6 well plates for RT-qPCR. For growth curve and cell count, myoblasts were plated at a density of 2x10^5^ cells/well in 24 well plates. At indicated time points, cells were harvested, and the number of viable cells was counted with a haemocytometer upon Trypan Blue staining. For proliferation assays, myoblasts were pulsed with 10 mM EdU (Invitrogen) for 2 hours immediately prior to fixation at indicated time point. Incorporated EdU was detected using the Click-iT EdU AlexaFluor Kit (Invitrogen) according to manufacturer’s instructions.

### Immunolabelling and Imaging

For immunolabelling, myoblasts were fixed in 4% paraformaldehyde/PBS for 10 min, washed in 3 x PBS for 5 min and permeabilised for 5 min with 0.5% triton X100/PBS. Subsequently, cells were blocked for 1 hour using 5% goat serum/PBS (blocking buffer). Primary antibodies were added in PBS and incubated overnight at 4C. Primary antibodies were: mouse monoclonal anti-PAX7 (Developmental Studies Hybridoma Bank; 1:500), chicken polyclonal anti-GFP (Abcam; ab13970; 1:1000), mouse monoclonal anti-MyHC (Developmental Studies Hybridoma Bank; MF20; 1:400) and rabbit polyclonal anti-MEF2C (CellSignaling; D80C1; 1:1000). Cells were then washed in 3 x PBS for 5 min, secondary antibodies added in blocking buffer and incubated for 1 hour at room temperature. Secondary antibodies were: Alexa Fluor 594 goat anti-mouse (Invitrogen; A11005; 1:1000), Alexa Fluor 488 goat anti-chicken IgY (H+L) (Invitrogen; A11039; 1:1000) and Alexa Fluor 647 goat anti-rabbit (Invitrogen; A21244; 1:1000). Nuclei were counterstained with 0.3 μM Hoechst 33342 (Invitrogen) in PBS for 10 minutes and mounted in PBS.

For staining and immunolabelling, muscle sections were washed 3x in Hanks’ balanced salt solution (HBSS) and stained with Alexa Fluor 647 conjugated wheat germ agglutinin (WGA) (ThermoFisher) diluted 1/50 in HBSS for 10 minutes at room temperature (RT). Sections were then permeabilized in 0.5% Triton/PBS for 10 minutes at room temp and then blocked in 5% goat serum/2% BSA/0.1% Triton/PBS for 1 hour. Sections were incubated in anti-PAX7 (DSHB) diluted 1:5 in blocking solution at 4° overnight, washed and then incubated in goat anti-mouse Alexa Fluor 594 at 1:400 in blocking solution at room temperature for 1 hour. Nuclei were stained for 10 min at RT in 1:1000 Hoechst 33342 (ThermoFisher) and mounted in mounting medium (Ibidi). Entire sections were imaged on a Zeiss AxioScan Z1 Automated Slide Scanner at 20× magnification. The number of PAX7-expressing nuclei over total number of nuclei in muscle biopsy was quantified using ImageJ. Cells were imaged using the EVOS™ M5000 Imaging System (Invitrogen).

### Mitochondrial Reactive Oxygen Species measurement

For mitoROS quantification proliferating myoblasts were plated at a density of 2.5x10^4^ in black, clear-bottom polystyrene 96-well plates (Nunc) and assayed 24 hours later. All fluorescent probes were applied in serum- and supplement-free Skeletal Muscle Cell Growth medium (PromoCell) for 25 min in the dark. Prior to mitoROS assaying, cells were washed twice with 1x HBSS (with Ca^2+^ and Mg^2+^; Sigma Aldrich), followed by incubation with 5μM mitoROS probe (MitoTracker Red CM-H_2_XROS, Sigma Aldrich) and 250nM MitoTracker Deep Red (Sigma Aldrich) to assay mitochondrial content for normalisation. For normalisation to cell input, DNA quantitation was performed by simultaneous incubation with Hoechst 33342 (0.5 μg/mL). After incubation with probes, cells were washed twice with 1xHBSS and fluorescence intensity was measured on a ClarioStar microplate reader (BMG Labtech) in spectral well averaging scan mode (100 flashes per well, scan diameter 6 mm). mitoROS probe fluorescence was normalised to mitochondrial content (MitoTracker Deep Red fluorescence) after normalisation to input cell quantity (Hoechst 33342 fluorescence), as simultaneously assessed via the respective fluorescence intensities in the same well and presented as fold change (mean ±SD) in PAX7mut compared to PAX7wt.

### RNA Extraction, Reverse Transcription and Quantitative PCR

Total RNA was extracted using the RNeasy kit (Qiagen) and quantified using a NanoDrop before being retrotranscribed with SuperScript III/IV First-Strand Synthesis System (Thermo Scientific). RT-qPCR was carried out using Takyon Low ROX SYBR 2X MasterMix blue dTTP (Takyon) as per manufacturer’s instructions on a ViiA7 thermal cycler (Applied Biosystems). RT-qPCR analyses were performed as previously described[29, 52]. Ct values of all genes analysed were normalised to the geometrical mean of Ct values of two housekeeping genes (e*GFP* and *RPLP0*) and fold changes were calculated using the ΔΔCt method (Livak and Schmittgen, 2001). Results are presented as mean value ± SEM of fold changes from independent experiments as indicated. Primers were previously described[29, 87, 88] or purchased from Sigma Aldrich and sequences are reported in Supplementary Table 4.

## Data analysis

Three images were taken per well at 10x magnification for each replicate and used for analysis in ImageJ (NIH, www.Fiji.sc). For immunolabelling, data is presented as the mean proportion of total Hoechst-positive cells ± SEM, N=3/4 biological replicates. For RT-qPCR, data was presented as average relative expression ± SEM, N=3. Data presented as mean±SEM from N=3/4 independently treated wells, considered biological independent replicates, from a representative experiment(s). Statistical significance was calculated in GraphPad Prism using unpaired t-test.

Alignment of PAX7 proteins across species was performed using Clustal Omega (ebi.ac.uk/Tool/msa/clustalo) with selected sequences retrieved from Ensembl (ensemble.org) and UniProt (uniport.org) databeses. Sequence identifiers are: Human PAX7 (*Homo sapiens*: P23759); Bonobo (*Pan paniscus*: A0A2R9AC01); Pig (*Sus scrofa*: A0A287A6Q7); Rat (*Rattus norvegicus*: D3ZRA8); Platypus (*Ornithorhynchus anatinus*: A0A6I8NAG5); Elephant (*Loxodonta africana*: G3UB47); Zebrafish (*Danio rerio*: C0M005). Gene Ontological (GO) analysis was performed on the two sets separately (Up or Down -regulated genes) (Supplementary Table 2) using g:Profiler (biit.cs.ut.ee/gprofiler) setting significant threshold to *p*=0.01 with Benjamini-Hochberg False Discovery Rate (FDR). Top 7 seven GOs in ‘Reactome’ and ‘Cellular Components’ were selected for visualisation. Heatmaps were created using Morpheus (broadinstitute.org/morpheus) and applying ‘Zscore’ adjustment to TPM (Transcripts Per kilobase Million) values of selected genes included in ‘Reactome’ GOs referring to RNA splicing and processing (R-HSA-72172, R-HSA- 8953854, R-HSA-72203, R-HSA-72163). Morpheus hierarchical clustering was applied blindly to assess the goodness of the genes (Figure 4C) to cluster separately proband sample biopsies from the three controls based on expression pattern of selected geneset.

## Data availability

All the raw data can be accessed on request.

## Compliance with Ethical Standards

The authors declare that the research was conducted in the absence of any commercial or financial relationships that could be construed as a potential conflict of interest. This study was approved by the Ethical Committee of the Helsinki University Hospital (HUS/16896/2022) and Ethics Committee of Santa Lucia Foundation IRCCS (CE/2022_020 approved on June 1, 2022). Written informed consent was obtained from all subjects.

## Author Contributions

MG and PSZ conceived, designed and planned experiments. MG generated the cell models and with PH and LMcG conducted experiments, acquired and analysed data. ENE generated and analysed data on muscle biopsies. PSZ secured funds for cell model generation and analysis. GFDN performed modelling of PAX7 structure. CS and EG performed genetic testing, methylation assessment and interpretation of results on proband and family members. MS and MJ performed variants analysis, RNA-sequencing processing and data analysis on patient biopsies. AB and VM isolated and generated the AB1190 human parental myoblast cell line. SB, ET, MM, AS, ER and GT acquired and analysed clinical data. MG, EG, PSZ and GT designed and conceptualized the study. MG, PSZ and GT drafted the manuscript. All authors reviewed the content for intellectual comment.

## Acknowledgments

We would like to thank the patient and his family for their participation in this study. We thank CSC – IT Center for Science, Finland for providing computational resources for RNAseq data analysis.

---

Supplementary Figure 1. **Genetic tests not confirming FSHD in the proband**

**A.** Schematic of Optical Genome Mapping (OGM) results on the proband showing the haplotype and the *D4Z4* alleles, excluding in-cis *D4Z4* duplications and mosaicisms.

**B.** Family pedigree showing 4qA/4qB haplotype in proband and family members.

**C.** Quantification of methylation levels (%) in proband for FSHD1 (DUX4-PAS; CpG.6 and CpG.3) and FSHD2 (DR1; CpG.1 and CpG.22) markers. Average methylation across general FSHD population (4qA/4qA) at specific markers is reported for comparison.

Supplementary Figure 2. **Transcriptomic analysis suggests no activation of compensatory pathways for muscle regeneration in proband muscle.**

Transcriptomic analysis shows no significant changes in indicated genes in proband muscle biopsies compared to controls.

Supplementary Table 1. **List of genes assessed on WES.**

Supplementary Table 2. **Gene Ontology (GO) analysis on DEG.**

Supplementary Table 3. **List of PAX7 variants reported with muscular or non-muscular phenotypes.**

Supplementary Table 4. **List of primers used in this study.**

