## Supplementary figures and images for "Biallelic *PAX7* variants cause a novel Satellite Cell-opathy with progressive muscle involvement resembling facioscapulohumeral muscular dystrophy"

### Supplementary Figure 1

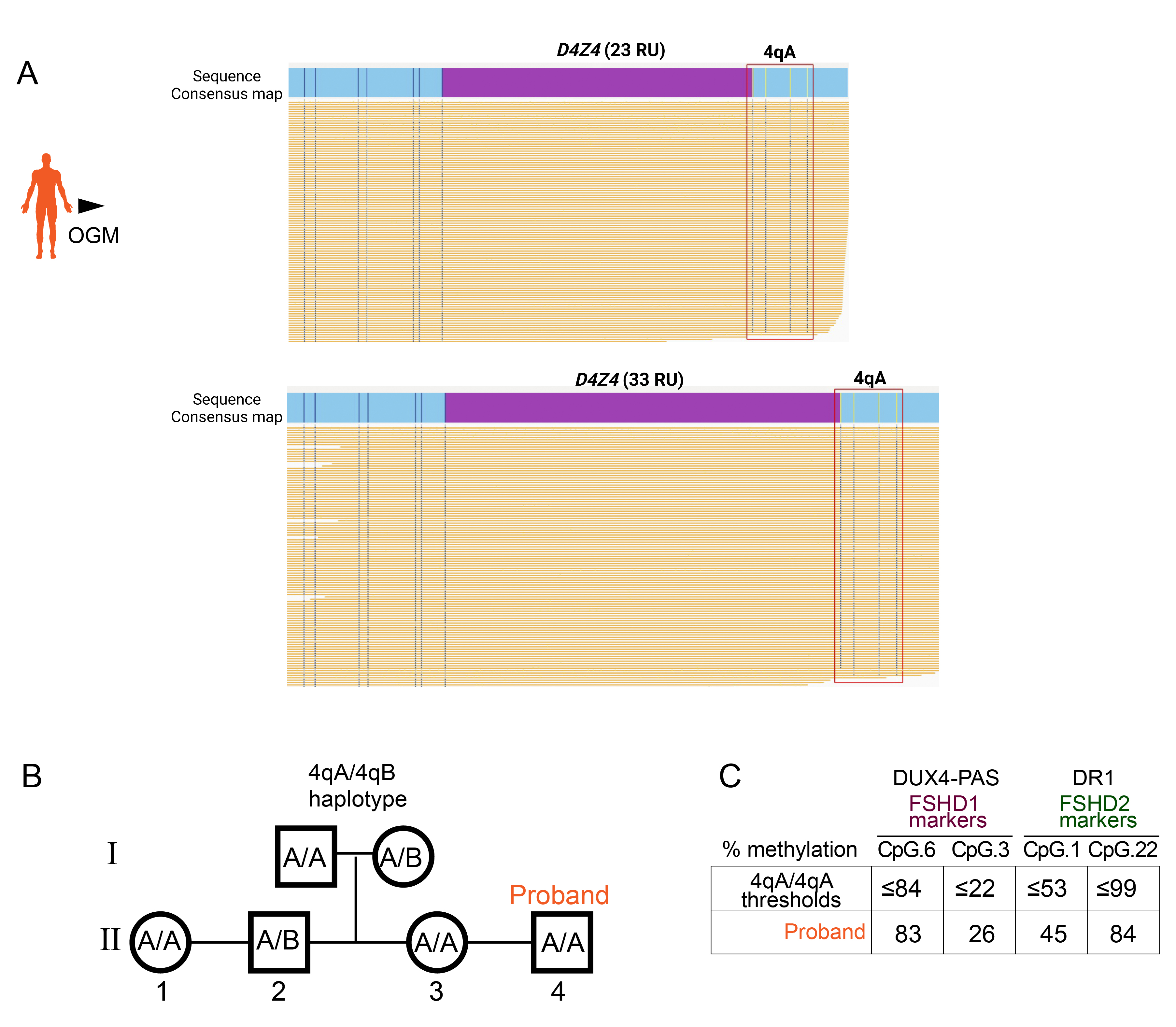

### Supplementary Figure 2

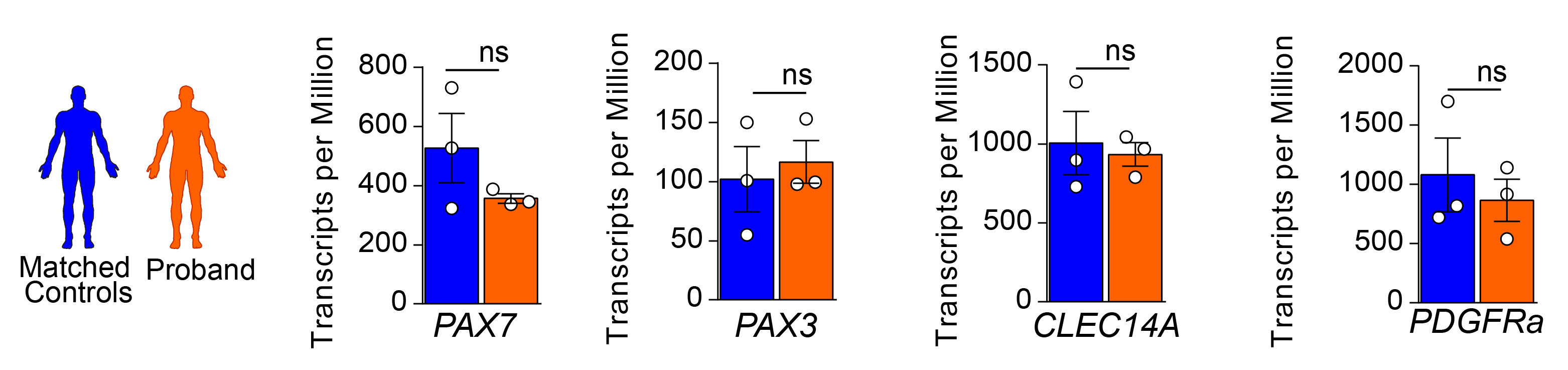
